# Modelling Singapore COVID-19 pandemic with a SEIR multiplex network model

**DOI:** 10.1101/2020.05.31.20118372

**Authors:** N. N. Chung, L. Y. Chew

## Abstract

In this paper, we have implemented a large-scale agent-based model to study the outbreak of coronavirus infectious diseases (COVID-19) in Singapore, taking into account complex human interaction pattern. In particular, the concept of multiplex network is utilized to differentiate between social interactions that happen in households and workplaces. In addition, weak interactions among crowds, transient interactions within social gatherings, and dense human contact between foreign workers in dormitories are also taken into consideration. Such a categorization in terms of a multiplex of social network connections together with the Susceptible-Exposed-Infectious-Removed (SEIR) epidemic model have enabled a more precise study of the feasibility and efficacy of control measures such as social distancing, work from home, and lockdown, at different moments and stages of the pandemics.

## I. INTRODUCTION

The 2019 coronavirus outbreak has become a pandemic. For the first time in human history, we observe a global concerted effort that tries to stem the spread of the epidemic with the semi-closing of national borders and grounding of international flights. Each country has its unique strategies to contain the virus in their territories. This ranges from complete lockdown in China and the laissez-faire approach in Sweden. While the most direct approach to arrest the virus is to create a vaccine against it, the lead time to vaccine development is long relative to the virulence of the disease that the best short-term strategy is social exclusion.

The coronavirus spreads through the simple mechanism of human-to-human contact. This is the reason why governments around the world implement lockdown and restrict social interactions. Ideally, if we could isolate everyone, there would be no room for the virus to survive. But this is socially impossible for family members. In addition, key economic activities still need to go on for the whole society to function, and this requires social interactions from workers providing essential services. There are two practical means to prevent the spreading of the virus through such a social route. One is to quarantine all infected individuals and also their contacts. This necessitates, however, all infected individuals to be identified immediately once they become infectious. But there is always a delay in such identification, and worst of all, some infected individuals are asymptomatic. The capability to carry out large-scale swap test has thus become important as it reduces not only the delay from disease onset to isolation, but also the number of undetected spreaders. The second approach is social distancing whose implementation has already severely disrupted the economies of many countries.

In order to properly manage the social-economic impact of the COVID-19 pandemic, governments would need a clear rationalization on the right policies to undertake. There are questions on whether lockdown should be implemented, like the case of Sweden versus its Scandinavian neighbors. And if lockdown were to be lifted, how gradual should we relax the restriction order? The means to address these questions together with the consequential advisories to the governments have been well served by mathematical and computational models. For example, a meta-population mobility model based on microscopic Markov chain approach [1] has been used to forecast the number of COVID-19 cases in each municipality of Spain. The outcome of the simulation had informed the Catalonian government ahead of time whether any Spanish region had exceeded its treatment capacity which led to a quick implementation of intervention measures that addressed the shortages. Meta-population based global epidemic and mobility model was also employed in another context to assess the efficacy of travel restrictions on the spread of COVID-19 at both the national and international level [2]. The results in this work showed that travel ban had mainly delayed the progression of the disease, with mitigation best achieved through transmission reduction intervention. In another modeling effort [3], the effects of reopening schools as part of exit strategies of lockdown was examined. It found that a gradual reopening of schools is necessary not to overwhelm the existing healthcare system or to cause a new second wave of epidemic transmission. All these models use real-world data to constantly calibrate their computations to drive the simulated scenarios close to reality and near to the ground truth. And their insights guide governments in their making of policies and decisions.

## II. MODEL

In this paper, we model the spreading of COVID-19 pandemics using the Susceptible-Exposed-Infectious-Removed (SEIR) model. Unlike the modified version of SEIR model used currently by a few research groups to simulate COVID-19 spreading [4, 5], our approach exploits both multiplex and temporal networks in conjunction with the SEIR model. Multiplex network had been used in Ref. [6] to study how the allocation of resources in a social support layer affects the spreading of virus in the physical contact layer. There are also prior research that combine SIR or SIS model with multiplex networks to investigate the effects of immunization [7], dynamical interactions between viral agents [8], emergence of epidemic phases [9], and contagion dynamics [10]. Recent research efforts on SEIR model on multiplex network have explored into the spread of rumors [11], and the transmission of COVID-19 in and between different layers of transportation networks [12].

The purpose and aim of our SEIR multiplex network model is to emulate the different forms of real-world social interactions in order to examine the effectiveness of various intervention strategies against the spread of COVID-19. The ability to tweak social interactions in mathematical models is crucial to find ways to curb the accelerating tides of epidemic spreading, in addition to giving insights on appropriate exit strategies from lockdown measures as the pandemic is entering the deceleration phase [13]. The main application of our model will be on the evaluation of the pandemic conditions in Singapore.

### A. SEIR Model

In our SEIR model, individuals are classified into four infection stages, namely susceptible, exposed, infectious, and removed. All individuals in the population are assumed to be susceptible to the virus before the pandemic begins. The model starts when the first infectious individual is imported into the population of susceptible individuals. Note that this imported individual is chosen at random from the population. Following this, *M*(*t*) individuals are selected uniformly at random at each simulated day *t* to model the *M*(*t*) number of imported cases. As the focus of our study is on the pandemic in Singapore, the information of these imported cases was obtained from the Singapore Ministry of Health webpage [14].

After a susceptible individual comes into contact with an infectious individual, the susceptible individual becomes exposed with a probability *p*. Note that it takes *T_e_* days on average before an exposed individual becomes infectious. Here, the exposure period is assumed to be Poisson distributed with a mean of *T_e_* days. Once the status of an individual is changed to infectious, it spreads the virus to each of its susceptible contact with a probability of *p*. Note that each individual can become infectious for different number of days depending on when they show the symptoms of infection and the time span from disease onset to social isolation. In addition, the infectious status of an otherwise asymptomatic individual is assumed to cease only when the individual has recovered. The infectious period is thus assumed to be gamma-distributed with a mean of *T_i_* days. Once recovered, an individual is no longer susceptible to the disease and can no longer become infectious.

### B. Social interactions

Real-world social interaction is often too complex to be represented by ideal complex network models such as the Erdőos-Renyí random network and the scale-free network. Here, we build a multiplex network [15] which is composed of multiple overlapping networks that describe the various types of social connections between agents to study the dynamics of the epidemic outbreak. Specifically, our multiplex network consists of a household network, a dormitory network, a workplace network, a temporal crowd network, and a temporal social gathering network. Note that this list of networks is not exhaustive. In principle, any community which is socially connected in a significantly different way and is not in the existing list of communities in the model should be added as a separate layer in the multiplex network. For example, a slum network will be included in our model for cities with a slum neighborhood.

Let us now go into the details of each of our social network. We have a household network in our model which captures the social interactions among family members within a household. It is represented by a complex network with a community structure. Agents that belong to the same household (or community) are more densely connected internally than with the other agents within the household network. The size of the household is assumed to be Poisson distributed with an average of *S_h_* members. On the other hand, social interactions within worker dormitories is modelled separately by a dormitory network as dormitories house a much larger number of residents compared to normal households. In addition, individuals within dormitories possess a larger number of social connections with other residents who stay in the same dormitory. Note that our special interest in worker dormitories stems from the occurrence of a virulent spread of COVID-19 within the dormitory clusters within Singapore.

We model the social contact within and between workplaces by a workplace network. In Singapore, 40% of the population are employed and work in a workplace. Each workplace is modelled as a community in the workplace network. The size of a workplace is assumed, however, to be gamma-distributed so as to include workplaces of extremely large size. Note that for the sake of simplicity, we have modeled social interactions within schools in the same way as that of a workplace.

The social interactions within public spaces, such as the public transportation system and markets, is very different from fixed households or in the workplaces. It involves short-term interactions among random groups of individuals each day. In our model, we select *f_c_* groups of *N_c_* agents on average uniformly and randomly on each simulated day to form *f_c_* fully-connected temporal networks. The spreading probability *p_c_* within each of these crowds of agents is assumed to be smaller than those of the household or workplace networks. Specifically, it is *p_c_* = 0.1*p*. In addition, we model social gatherings such as religious services, academic conferences, and large-scale dinner events in a similar fashion albeit with a different topology for their social interactions. On each simulated day, *f_g_* groups with an average size of *N_g_* agents are formed uniformly at random with the topology of scale-free temporal networks of average degree *k_g_* as their mode of transient social interactions.

## III. METHODS AND PARAMETERS

### A. Reproduction number

The reproduction number *R*_0_, which is defined as the number of secondary infections caused by an infected individual, is a closely monitored measure that reveals the current state of the epidemic outbreak. It serves to inform government officials and epidemiologists who are tasked to contain the pandemic whether an outbreak is imminent. If the reproduction number is above 1, the infectious cases increase and the occurrence of an outbreak is expected. On the other hand, if *R*_0_ is below 1, the number of infectious cases diminishes and the epidemic is well-contained. *R*_0_ changes dynamically as the virus spreads to different parts of the population in consequence of social interactions among individuals and communities. The introduction of interventions such as travel ban and social distancing have the positive effect of lowering the value of *R*_0_. Indeed, the reproduction number was found to decline from 2.35 to 1.05 after Wuhan was locked down in late January [16].

Two common methods to estimate the reproduction number are the maximum likelihood estimator and the branching process estimator [17]. Here, we adopt the branching process estimator to evaluate *R*_0_. First, we group a time series of daily number of new cases *n* = {*n*_0_*, n*_1_*, n*_2_*, · · ·*, *n_T_*} into *m* generations, where each generation consists of *T_e_* number of data points. We then form the set:

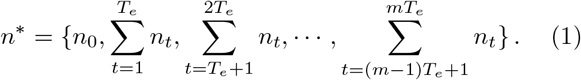

Note that the mean exposed time *T_e_* gives the time between infections in consecutive generations or the serial interval. As we are concerned with an evolving *R*_0_, we estimate *R*_0_ for each generation *i* as:

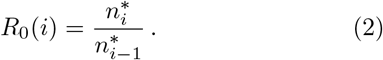

New cases are reported and included in an epidemic database after the infected individuals are tested positive. There is however no simple or direct relationship between the number of reported cases and the number of actual new infectious cases. In Ref. [18], a linear regression model was used to impute the missing onset date of infection taking into account the reporting delay distribution. Here, we deduce the onset date with our SEIR model. Specifically, we simulate *M* = 50 realizations of the pandemic with our SEIR model and select 20% of them which best-fit the epidemic curve under study. The number of new infectious cases are then deduced from these realizations.

### B. SEIR model with multiplex networks

In our simulation, we use an effective population size of *N* = 1, 000, 000 instead of the actual Singapore’s population size of 5.6 million in 2020. This assumption is reasonable because the number of infected cases 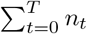 is relatively small compared to the actual population size. Hence, it is feasible to assume that a large proportion of the population has a vanishing chance of coming into contact with any infectious individual during the period under study. This assumption has the advantage of reducing the time of the simulation.

In a similar vein, we have assumed that 55, 000 foreign workers stay in the dormitories instead of the actual number of 320, 000. For simplicity, we have also assumed that each dormitory accommodates 1, 000 individuals. As a result, our dormitory network shall consist of 55 communities. We have set the average degree of the dormitory network to be *k_d_* = 60.15. Most of the network connections are within the community of a dormitory, with each individual in a dormitory connected to about 0.15 other individual in the other dormitory.

The rest of the population of size 945, 000 are assumed to stay in households of average size *S_h_* = 2.5. A sample household size is drawn randomly from a Poisson distribution with a mean of *λ* = 2.5. Members in the same household are socially connected with a probability of 0.95. Moreover, we assume that on average each individual in the household network is socially connected to 0.4 other individuals in another household.

Next, we model economically active individuals in the household network by connecting 45% of them within the workplace network. Note that these individuals include students “working” in schools. Also, 90% of the individuals in the dormitory network are included in the workplace network. Furthermore, workers who stay in dormitories have higher chance to work in the same workplace. The workplace network is assumed to be modular in nature with an average community size of *S_w_* = 4. The size of a workplace is drawn randomly from a gamma distribution with a shape parameter of 2 and a scale parameter of 2. Members within the same workplace are socially connected with a probability of 0.7, while the social connection between workers of different workplace is set at 0.08. In addition, we have introduced a small number of inter-network household-to-workplace links that connect nonworking household individuals with workplace individuals. Similar household-to-dormitory inter-network links are also inserted within our multiplex network model.

In the case of our temporal crowd network, we have set *f_c_* = 500 and *N_c_* = 50 for each simulated day. The parameters for the temporal social gathering network are: *f_g_* = 200, *N_g_* = 50 and *k_g_* = 8.

Our simulation begins with the scenario in 21 Jan 2020, two days before the first reported case of COVID-19 in Singapore, and end on 13 May 2020. Singapore implemented Circuit Breaker (CB) on 7 April 2020 to restrict social interactions among its residents. After the CB, we set *f_g_* = 0 and *f_c_* = 200. We turn off social connections in 85% of the workplaces, with the remaining 15% continue to be functional with essential workers physically at work. We have also turned off social connections between workplaces. On the other hand, social connections within the households remain unchanged, albeit we turn off 95% of the social connections between households while maintaining 5% inter-household connections to represent violations of stay-home measures. To model Singapore’s government effort in revamping the living conditions of the foreign workers, we reduce the social connections within dormitories by cutting down 25% of the connections every 5 days until there are 25% of the connections left. Finally, we set *p* to 0.1 [19]. *T_i_* is set to 3 for the first 30 days from 21 Jan 2020 to reflect the longer delay from disease onset to case report at the early phase of the pandemic. It is adjusted to a value of 2 afterward. The values of the parameters are either judiciously drawn from the literature or they are calibrated from the real-data.

## IV. RESULTS AND DISCUSSION

The outcome of our simulation is illustrated in Fig. 1. The simulation results with CB show good correspondence with the real data for both the total number of reported cases as well as the reported cases for just the community spread of COVID-19 (i.e., cases that exclude infections within dormitories). In both cases, we observe signs of a start of a flattening of the epidemic curve. Our results here is consistent with that of [20] which predicts that Singapore is near to the theoretical ending of the pandemic based on predictive modeling.

**FIG. 1.**
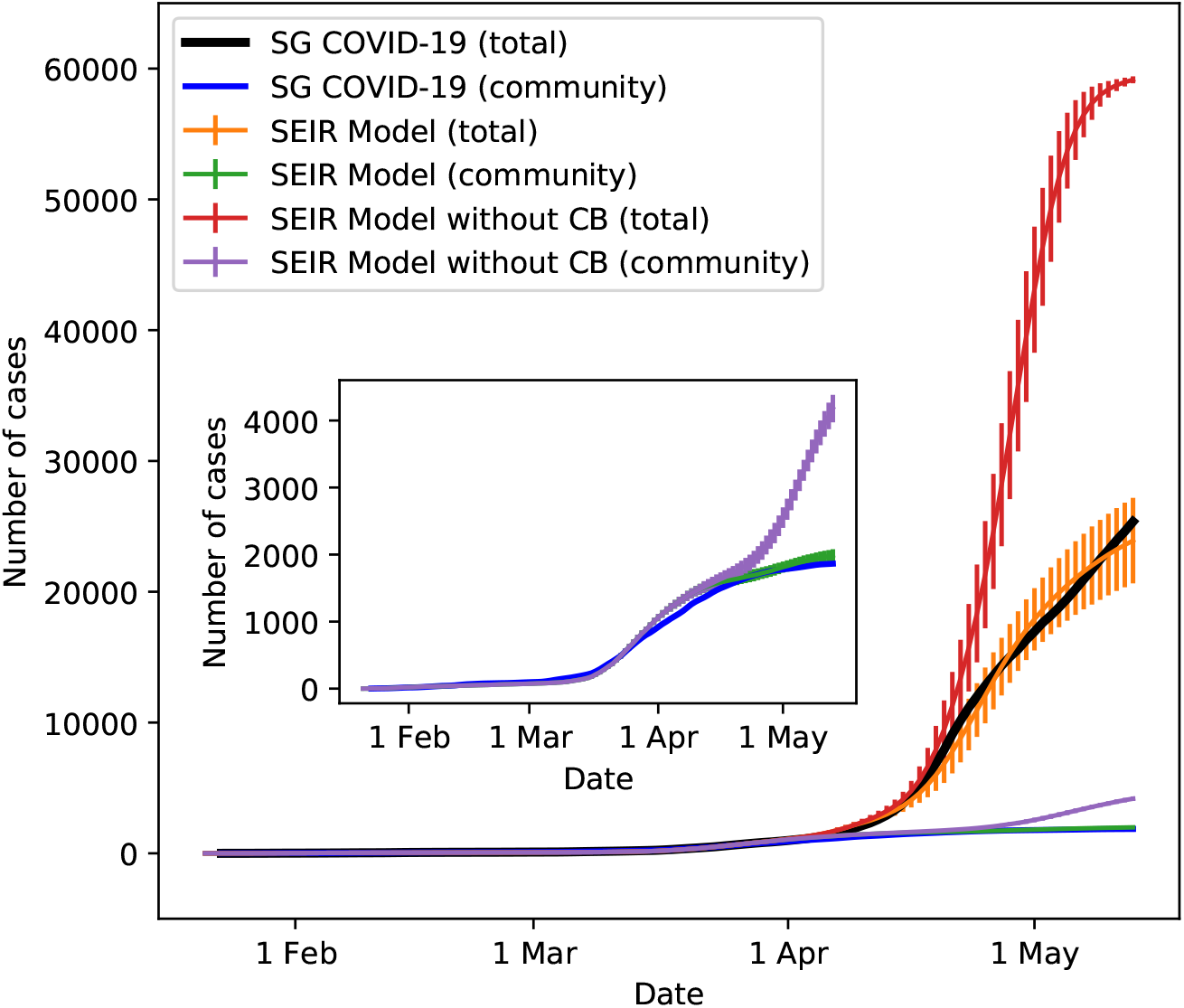
Epidemic curves for the spreading of COVID-19 in Singapore. The curves are shown for the real data, and for the simulated dynamics with and without implementation of the Circuit Breaker (CB) measures. The inset shows an enlarged view of the spreading of COVID-19 within Singapore’s community. Standard deviation is shown as vertical line for the simulated dynamics.

Our model allows us to simulate the scenario if the government has not implemented the circuit breaker and has not revamped the living conditions of the foreign workers. Under this scenario, our results show an exponential rise in the total number of cases. Note that the number of cases will be even more if not for the maximum capacity of 55, 000 dormitory workers we have fixed in our simulation. This explains the appearance of a saturation in the curve for the total number of cases without CB. Thus, the CB has successfully contained by much more than 3 times the spread of COVID-19, and it has correspondingly reduced and prevented the stress on the medical facilities due to the pandemic.

Figure 2 illustrates the progression of the pandemic in Singapore through the dynamics of *R*_0_. We observe that *R*_0_ predicts the emergence of three waves of enhanced COVID-19 transmission when *R*_0_ becomes larger than one. Moreover, each successive wave is observed to have a greater transmission impact compared to the preceding one. The first wave happened on 9 Feb 2020 according to Figure 2. It is the precursor to an infection cluster at the church of the Grace Assembly of God with its first reported case on 12 Feb 2020 (which is also linked to reported cases from the Life Church and Missions Singapore) [21]. There are a total of 33 infected individuals in this cluster. The second wave began on 23 Feb 2020 (see Figure 2). It is related to another infection cluster due to a private function at SAFRA Jurong with its first reported case on 27 Feb 2020. The total number of infected individuals for this cluster is 48. These two waves of COVID-19 transmission are in fact exceedingly small relative to the third wave which was started on 29 Mar 2020 based on the *R*_0_ illustrated in Figure 2. The third wave is a very large wave caused by the circulation of COVID-19 within the dormitories of the foreign workers. Although the first reported case was published on 5 April 2020, its presence could already be preempted from the reported case of infection within the construction site in the Raffles Place area on 3 April 2020. The total number of infected foreign workers in the dormitories is about 23, 000 by current count, and the cluster is still growing. The time delay between the initiation of each wave and the first reported case is consistent with observations of a delay between onset of symptoms and the diagnosis for COVID-19 [21].

**FIG. 2.**
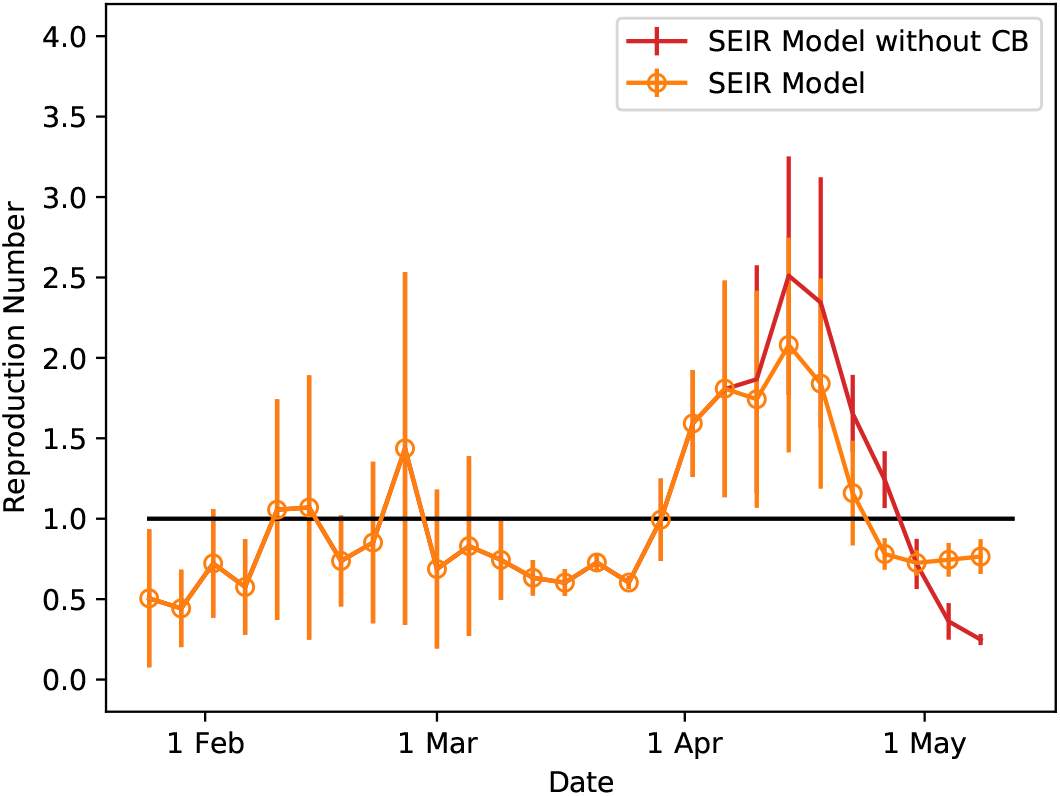
The dynamical evolution of the reproduction number *R*_0_ derived from the SEIR multiplex model with and without Circuit Breaker (CB) measures.

Without CB, we expect the transmissibility of COVID-19 to be higher, which is confirmed by the results in Fig. 2. However, the *R*_0_ should continue to rise beyond the middle of April 2020 if not for the maximum capacity imposed on the dormitory workers by our model, as explained earlier with regards to the epidemic curve without CB.

## V. FINAL REMARKS AND CONCLUSION

In this paper, we have employed a SEIR multiplex network model to simulate the progression of the COVID-19 pandemic in Singapore. We have demonstrated the utility of our model in evaluating the efficacy of Circuit Breaker in containing the spread of the virus. With the Circuit Breaker approaching its end on 1 June 2020, there are uncertainties on the strategic approaches to lift the various imposed social restriction orders. We envisage that our SEIR multiplex network model could play the role of simulating potential strategies with differing level of social interaction limitations, with the purpose of determining the right amount of social-economic functions that can be restarted in Singapore without causing any harm to the successful containment effort already achieved on COVID-19.

As such, we have explored the use of our model to simulate a gradual relaxation of social restrictions from 1 June 2020 to 30 June 2020. Here, we assume a full restoration of inter-household social connections while intra-household connections remain unchanged as in CB. Furthermore, we consider 30% of the non-essential workplaces to become socially connected again within the workplace network, with 15% of workplaces for essential services to remain status quo. As a result of the returning of the workforce to their workplaces, we increase the crowd frequency to *f_c_* = 300. We also increase *f_g_* = 200, but with a smaller average group size of *N_g_* = 25. Finally, we assume the social connection within the dormitory network to maintain at 25%.

Our simulation results based on these parameters (see Figs. 3 and 4) show that the strategy of gradual lifting of social restrictions will continue to flatten the epidemic curve, while a drastic resumption of social interactions to the period of 21 Jan 2020 will lead to another large wave of COVID-19 transmission. Hence, our model indicates that prudent public health policies that avoid massive social interactions are quintessential in maintaining the current state of pandemic control.

**FIG. 3.**
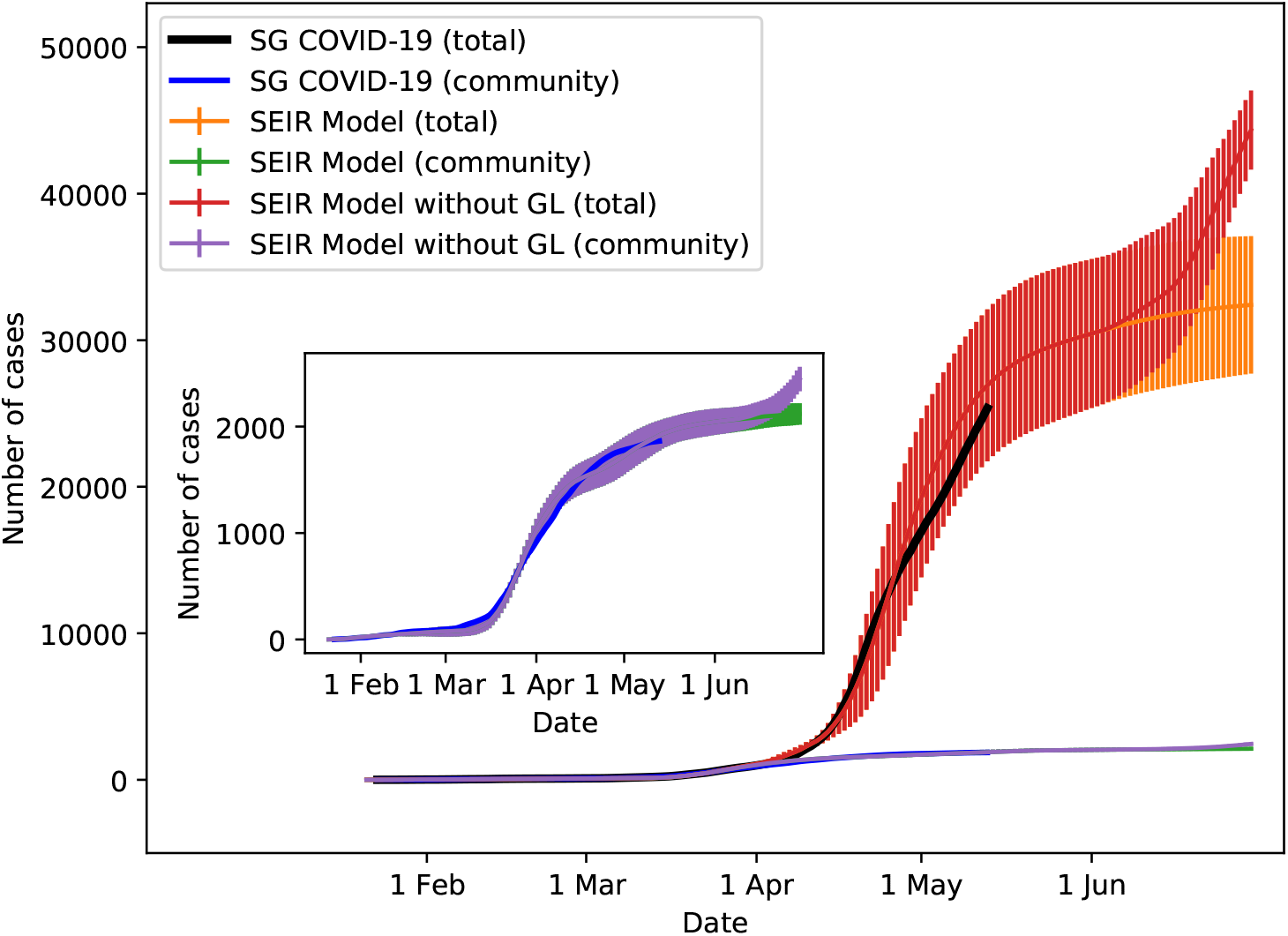
Epidemic curves for the spreading of COVID-19 in Singapore. The curves are shown for the real data, and for the simulated dynamics with and without implementation of the Gradual Lifting (GL) measures. The inset shows an enlarged view of the spreading of COVID-19 within Singapore’s community. Standard deviation is shown as vertical line for the simulated dynamics.

**FIG. 4.**
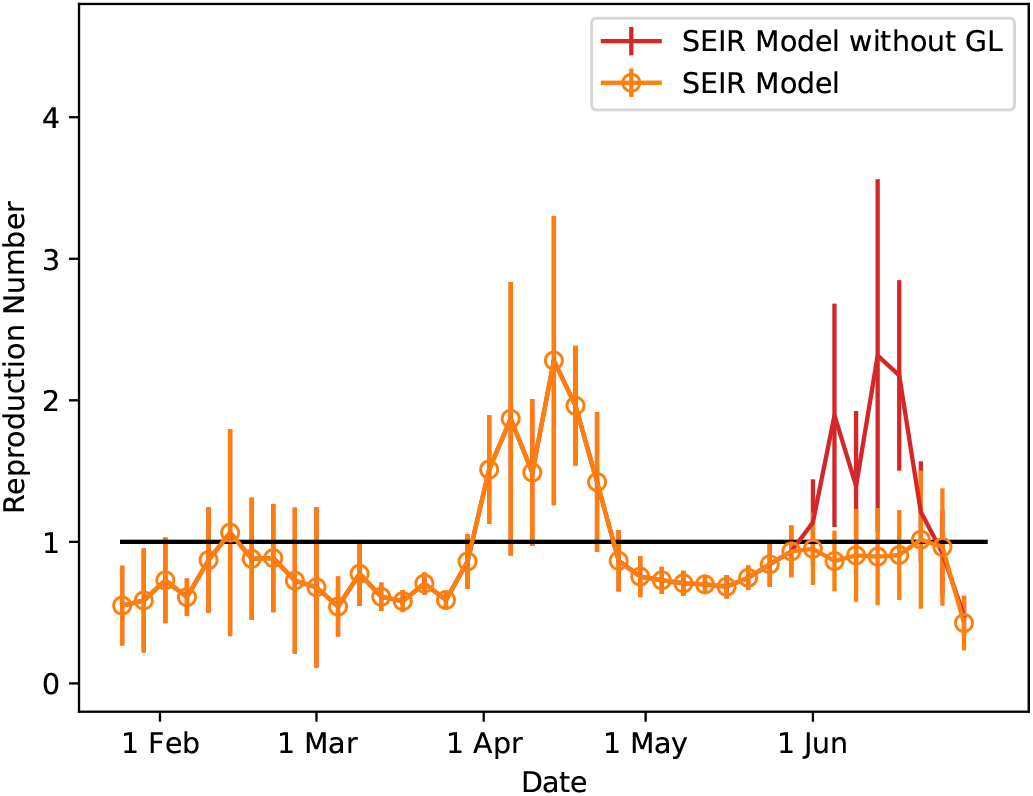
The dynamical evolution of the reproduction number *R*_0_ derived from the SEIR multiplex model with and without Gradual Lifting (GL) measures. Note that the two very small waves of COVID-19 transmission at 9 Feb 2020 and 23 Feb 2020 of Fig. 2 reported in the main text are absent in this plot due to the large statistical fluctuations.

For future work, we plan to scale up our model to simulate an effective population size of 2, 000, 000. With the increase of the effective population size from 1 million to 2 million, the members in each network will also double, i.e. the size of dormitory becomes 110,000, while that of household becomes 1,890,000, for the next version of our SEIR multiplex network model.

## Data Availability

Data is available at Singapore Ministry of Health webpage - https://www.moh.gov.sg/covid-19

